# Health literacy, information access and COVID-19 vaccination hesitancy among foreign-born persons in Sweden – a focus group interview-study

**DOI:** 10.1101/2023.12.27.23300586

**Authors:** Mia Söderberg, Juhaina Swaid, Kristina Aurelius, Annika Rosengren, Kristina Jakobsson, Maria Magnusson

## Abstract

**Background:** Lower rates of COVID-19 vaccination have been observed in individuals with an immigrant background, yet if this relates to barriers to obtaining reliable information is unknown. This exploratory interview study investigated health literacy and information access as determinants for vaccination hesitancy towards the COVID-19 vaccine among foreign-born individuals in Sweden.

**Methods and findings:** We used purposive sampling to recruit foreign-born adults from low- and middle-income countries and health guides and doulas who were assigned to spread COVID-19 related information in immigrant-dense urban areas. Data were collected using semi-structured focus group interviews, which were transcribed verbatim and analysed according to systematic text condensation.

Ten participants were included who were gainfully employed as health guides/doulas, or in other jobs, full-time students, or housewives. Four main themes emerged: 1) Limited health literacy, 2) Consequences of not using official Swedish information, 3) Decision-making on COVID-19 vaccination, and 4) Suggestions to improve information dissemination effectiveness. The lack of health literacy in official institutions, health care personnel and recipients alike led to little use of official information. Instead, most participants relied on social media, social contacts and international media, through which a lot of contradictive and negative information about the vaccine was spread. The decision to get vaccinated or not was a process fraught with insecurities about the effectiveness and side effects of the vaccine, which was balanced against wishing to be protected and contributing to the battle against COVID-19. Suggestions for information dissemination improvements from the participants were to produce multilingual information and to increase the use of transmission through social interaction with trusted persons and platforms.

**Conclusions:** An inadequately adapted information outreach prevented some members of the society from making fact-based decisions about getting vaccinated. Several suggestions for improving dissemination were brought forth that can be tested in future communication strategies.

**Author summary:** *Why was this study done?:* People with an immigrant background have consistently displayed a lower vaccination uptake than the general population. This study investigated aspects of health literacy, information access and vaccination hesitancy in foreign-born individuals in Sweden.

*What did the researchers do and find?:* Few participants accessed official information about the COVID-19 vaccine, mainly because of poorly adapted information outreach, language barriers and not knowing Swedish institutions. Instead, they turned to a multitude of other sources from which conflicting and inaccurate information was spread, lowering their confidence in the COVID-19 vaccine.

*What do these findings mean?:* In the case of national emergencies, important public health information does not reach everyone equally, obstructing the possibility for some to make an information-based decision on how to protect their health.

## Introduction

For widespread infectious diseases, vaccination is one of the most effective public health interventions to slow down and prevent severe disease development and death. When the new SARS-CoV-2 virus rapidly developed into a pandemic (1, 2), large efforts were immediately made to develop a vaccine. In December 2020, the first COVID-vaccine BioNTechs produced by Pfizer was approved by the European Medicines Agency and shortly thereafter Astra Zeneca’s vaccine Vaxzevria and Spikevax Moderna were also approved. In Sweden, the first vaccine batches were distributed in December 2020, aimed at-risk groups such as those living in long-term care facilities, adults aged 70 years and older, and healthcare staff who came in contact with potentially COVID-19-infected patients (3). Gradually more subpopulations became eligible and in early June 2021, everyone aged ≥18 years was included in the vaccination programme. The vaccination strategy was deemed effective as 75% of all elderly in long-term care facilities had received a first dose in February 2021 and by mid-July, 70% of the adult Swedish population had received at least one dose (3).

The success of any vaccination programme depends on people’s willingness to receive the vaccination; however, this tends to vary between different societal groups. As reported by the Public Health Agency of Sweden, in April 2021, 91% of all Swedish-born people aged *≥*80 years had received their first dose, while a notably lower coverage was observed in same-aged persons from South America (66%), Middle East (62%), North Africa (59%) and Africa, Other (44%) (4). Similarly, in a general population cohort study (n= 972,723) with national register data extracted in May 2021, immigrants from low- and middle-income countries displayed a fourfold risk of not being vaccinated compared to Swedish-born persons, adjusted for age, sex, income, country of birth, and household composition (5). Based on the observations of low vaccination coverage in some groups, targeted campaigns started, but the low vaccination uptake persisted. In another register study with data on sex, occupation, country of birth and vaccination status until November 2022, men and women in white-collar occupations born in a Nordic country displayed a vaccination coverage of 80-90% (6). The corresponding rates were markedly lower in non-Nordic white-collar men (60-80%) and women (65-85%), and even more so in blue-collar workers born outside the Nordic countries (men: 30-70%; women: 45-70%). Such findings are consistent with several studies done outside Sweden (7). Vaccination hesitancy is defined as a refusal or delay in getting vaccinated. The decision involves balancing potential benefits, perceived vulnerability, and the likelihood of harm, and can be further influenced by factors such as a sense of urgency and prior negative experiences (5). Furthermore, some aspects are specific to having an immigrant background. A review of qualitative studies on vaccination hesitancy in immigrants during the vaccination rollouts for COVID-19, HPV, influenza, hepatitis B, and pneumococcal infections, found several determining factors such as a lack of knowledge of the vaccine’s existence and function, misinformation, distrust of official institutions and cultural bias (8). Among these findings, a lack of knowledge of the existence of a vaccine and its main benefits could be considered especially alarming.

Immigrants’ lack of access to information from official institutions is a recurring theme (9, 10) and relates to a lack of “health literacy” (11, 12). Health literacy is a concept that reflects the ability to spread and obtain information that can promote and protect health. It involves the disseminator’s ability to adapt an information campaign for it to reach all members of a target group equally. Likewise, it involves the receptor’s ability to find, understand, and apply information that can enhance decision-making about health and disease prevention (13). In a recent interview study, language barriers and not knowing the existence and role of official public health and healthcare outlets contributed to a lack of access to COVID-19 information among foreign-born persons (14). In contrast, among students enrolled in the “Swedish for Immigrants” programme, to whom information on COVID-19 was spread through established channels and in several languages, the majority of the respondents reported that they had received adequate information, and were informed about protective actions and the current development of COVID-19 in Sweden (15).

Despite consistent evidence of less access to vital public health information and a lower vaccination uptake among immigrants, little is known the extent to which this affects the decision to accept a vaccine. A systematic review study from 2018, focusing on non-interactive communication, found that age and the country in which the vaccination program was conducted were the most decisive factors for accessing information about a vaccine (16), but information from immigrant groups was lacking. Another review study, covering several vaccination programmes including the SARS-CoV-2 vaccine rollout (17), merely found associations between health literacy and influenza vaccination uptake in persons aged 65 years and older. Out of 21 included studies, one investigated American Hispanic women, but failed to find any association between health literacy and attitudes to the influenza vaccination. This study did not specify if these women were foreign-born or born in America, which is relevant for language sufficiency and knowledge of local healthcare systems.

In Sweden, as well as internationally, vaccination hesitation among foreign-born persons has been observed despite extensive efforts to spread information about the existence and benefits of the vaccine. Nevertheless, little is known about the underlying reasons for not taking the vaccine, especially as told by people from this sociodemographic group themselves. This explorative qualitative interview study aims to investigate health literacy, information access and hesitancy towards taking the COVID-19 vaccine in foreign-born persons living in Sweden.

## Methods

### Recruitment of the participants

Participants were recruited during spring-summer 2022 by using purposive sampling, a common method when there is a clear idea of participant attributes that are of particular interest to the study aims. Thus, we included foreign-born adults from low- and middle-income countries, among whom we tried to seek out people who displayed limited Swedish language proficiency and expressed hesitancy towards taking the vaccine. We also recruited health guides and doula, who during the pandemic had an extended role in spreading COVID-related information in immigrant-dense suburbs. In Sweden, health guides serve as a bridge between foreign-born persons and Swedish community functions, with the assignment to facilitate communication between organisations and citizens. The aim is to help people promote their health and to ensure everyone in society has access to the same information. Doulas have a similar assignment but primarily provide guidance and emotional support to women during childbirth. All participants were recruited through the social networks of two health guides, who at the time were employed at the East Gothenburg Health Centre (Hälsoteket Östra Göteborg) and were contracted to recruit participants and conduct the interviews.

Ten participants were interviewed in three different focus groups: one Arabic-speaking group (two women, two men), one Somalian-speaking group (three women) and one group with health guides/doulas (three women). The participants’ age ranged from 20-60 years, and their countries of origin were Somalia, Syria, Lebanon, Iran, Iraq and Kurdistan. Out of the participants who were not employed as health guides/doulas, three were gainfully employed in other jobs (grocery store staff, assistant nurse and child nurse), two were enrolled in vocational training programmes, and two were housewives. All participants received information about the study both orally and written and signed a written consent form.

### Data collection

The interviews were conducted by Juhaina Swaid (JS) (second author), Saido Mohamed (SM) and Mia Söderberg (MS) (first author) according to a semi-structured methodology with a set of open-ended questions to encourage the participants to freely describe their thoughts around each subject. The interview guide comprised three main topics: 1) access and barriers to information on COVID-19 and the COVID-19 vaccine, 2) whether the obtained information was helpful in the decision to get vaccinated, and 3) how official information outreaches can become more effective. Both JS and SM have had prior contact with the participants through their work as health guides. MS had not encountered any of the participants before the interviews.

One of the focus group interviews was carried out in Arabic (by JS) and one in Somali (by SM), the native language of both the participants and interviewers in each group, while MS had an assisting role. The interview with health guides and doulas was carried out in Swedish by MS assisted by JS. Swedish was not the native language of the health guides/doulas, but MS and JS perceived the participants’ level of competence in Swedish as moderate to advanced, and bias due to language difficulties was assumed to be small. At one point a doula explained herself in English to make sure to convey her message properly, else the interview was conducted in Swedish. All interviews were recorded, interviews in Arabic and Somali were translated to Swedish by certified interpreters, and all interviews were then transcribed verbatim.

### Analyses

The main data analysis was conducted by first author MS and second author JS. The chosen analysis method was systematic text condensation as proposed by Malterud (18) which focuses on paying attention to the participants’ experiences and perspectives, while also trying to find and understand underlying processes. MS and JS first read the printed transcript of all the interviews separately, without singling out meaning-bearing information to get the overall perception of the contents of the data. The transcripts were then read again, but now to identify statements which contained meaningful information about the study objectives. This was done manually by marking out meaning-bearing units with a pencil or colour marking and adding comments in a Word document. In the next step, these units were revised and labelled into broad categories, re-read and divided into main- and sub-categories. MS and JS then compared their themes and categorisation of these themes, and further revised the sorting of these themes. The results were then presented to all the co-authors, and after discussion, two of the main themes were slightly altered.

## Results

Our analyses resulted in four overarching themes: 1) Limited health literacy, 2) Consequences of not using official Swedish information, 3) Decision-making on COVID-19 vaccination, and 4) Suggestions to improve information dissemination effectiveness (Table 1).

**Table 1.** Main themes and sub-themes.

| Main Theme | Sub-theme |  |
| --- | --- | --- |
| **1. Limited health literacy** | 1.1 In official health organisations | No information in other languages than Swedish<br>Ineffective dissemination channels |
|  | 1.2 In health care staff | Did not respect or understand information requests<br>Did not provide nuanced information |
|  | 1.3 In foreign-born recipients | Language barriers<br>Lack of knowledge of Swedish institutions |
| **2. Consequences of not using official Swedish information** | 2.1 Used other information sources than official Swedish sources | Social contacts<br>Social media<br>International TV and radio |
|  | 2.2 Domination of unreliable information | Difficult to evaluate trustworthiness<br>A large spread of disinformation with a negative focus<br>Rumours exacerbated by the local community |
| **3. Decision-making on COVID-19 vaccination** | 3.1 Aspects that increase vaccination hesitancy | Taking a risk without fully knowing the benefits or hazards<br>Is the same dose appropriate for all ages and ethnicities?<br>Is it safe for people with pre-existing diseases?<br>Do not think it is necessary |
|  | 3.2 Aspects that increase vaccination motivation | Understanding the protective functions of the vaccine<br>Want to fulfil a societal duty<br>Pressure from an employer or school<br>Want to enter other countries |
| **4. Suggestions to improve information dissemination effectiveness** | 4.1 Information in many languages and formats | Written and spoken information in several languages<br>Regularly information outreach through official channels |
|  | 4.2 Communication through social interaction | Facilitates two-way communication<br>Utilise trusted people and platforms<br>Periodical information meetings in person or via online platforms<br>Invite specialists to open meetings in the local areas |
|  | 4.3 Increased role of school, work and local healthcare centres | Systematic spread of information<br>More adherent to requests |

### 1. Limited health literacy

The interviews revealed several examples of limited health literacy among official institutions, healthcare workers and foreign-born recipients, which diminished the successful spread and uptake of information about the COVID-19 vaccine.

#### 1.1 In official health organisations

A lack of health literacy in official institutions manifested itself as inadequate information design and dissemination methods for people outside the majority society. The two main Swedish institutions for health information, the Swedish Public Health Agency and 1177, the health care system’s unified point of contact for health care advice were unknown to most of the participants Thus, information communicated through the web pages and social media accounts of these institutions did not reach them. Information was initially only published in Swedish and when it became available in other languages, some were incorrectly translated. Because of such shortcomings, these outlets were not known or trusted when the vaccine was introduced.

*“(Interviewer: How long did it take until they translated into more languages?) A long time, several months. (Interviewer: Several months, then maybe in May 2020?) After May.”*

*“I read [translated information] at the beginning of COVID, and it was translated incorrectly. And people were scared because of what was wrongly translated.”*

*“Many wonder, do the authorities care about us?”*

#### 1.2 In health care staff

Personnel at the local health care centres or at the vaccination sites, whom the respondents met in person, had an opportunity to be made aware of knowledge gaps and respond to them. Yet, the respondents expressed that the personnel seemed to neither care for nor would answer questions about safety or how a vaccine works. Many also feared that pre-existing diseases could interfere negatively with the vaccine, but such issues were not taken seriously, which made them feel like the personnel did not understand that they had many concerns about taking the vaccine and that addressing these issues could lessen their worries.

*“You heard that someone had died after the vaccination or that someone else was in a coma because they had received the vaccine when they already had a cold… I was afraid… I wanted to examine myself before the vaccination. But every time I was denied, and they just said that the vaccine was harmless, and I should take it.”*

*“Some young people had been vaccinated with it at the first dose and with a different kind of vaccine at the second dose… You wonder what was wrong with the first vaccine? And the only answer you got was that it [the vaccine] does not pose any danger.”*

#### 1.3 In foreign-born recipients

The most notable limitation of health literacy among the respondents was their limited Swedish proficiency, which prevented an understanding of information and made it difficult to navigate the Swedish health systems. These obstacles appeared to be less pronounced among participants who held a job, who mostly perceived Sweden’s information outreach as satisfactory. Some respondent speculated that newly arrived, elderly, and housewives had the least access and that information in many languages would have made a difference.

*“A large proportion of those who did not get vaccinated were immigrants. (Interviewer: Why did information not reach their [residential] areas?) Language deficits.”*

*“You have to use another language. That is my opinion. Many have a mother tongue other than Swedish… There are many mothers who do not know Swedish, and (you) have to explain a lot of Swedish.”*

*“I think the information was good, it was clear and in several languages… The Swedish authorities made no mistakes, they gave enough information.”*

### 2. Consequences of not using official Swedish information

In the absence of official Swedish information, or to seek out additional information, the respondents turned to personal contacts, social media or international TV and radio with broadcasts in their native language. This exposed them to a large flow of information from multiple sources that was difficult to verify and often dominated by negative aspects of the vaccine. A lot of misinformation then appeared to be circulated in the local community, contributing to the spread of rumours, confusion, and fear.

#### 2.1 Used other information sources than official Swedish sources

Information was gathered mainly from social contacts, social media, and international TV and radio channels in their native language. The use of social media (Facebook, YouTube and Instagram) was concentrated on platforms by people with a high social standing or who claimed to have a medical education. With time, the usage of official Swedish outlets appeared to increase but many, both the respondents and people they knew, continued to rely on unofficial outlets.

*“When they are new (in Sweden) and do not know the Swedish language, they use their mobile phone and check social media that are in their language. (Interviewer: Is that what they turn to the most?) Yes, because it is in their language.”*

*“I don’t work, I’m a housewife. There was a TV channel that published the number of deaths from COVID-19… It felt like psychological warfare. I was very scared… (Interviewer: How did you know that there was a COVID vaccine in Sweden?) I found out when my husband got vaccinated.”*

#### 2.2 Domination of unreliable information

The large amounts of information from multiple sources made the respondents feel like they were exposed to “a flood” of information whose trustworthiness was difficult to evaluate, and which mostly focused on negative aspects of the vaccine. For example, a lot of information in social media seemed to come from people who claimed to have a medical degree, but this could not be verified. This type of loosely based information was then circulated in their local community and exacerbated by rumours and personal stories about negative experiences.

*“(Interviewer: Do you think this was positive or negative, that people obtained information from other countries or googled it online?) It was more negative than positive. The information was conflicting, and some information was untrue. It was a contributing factor to fear among people.”*

*“I remember a video clip in which a doctor explained how to clean the home with chlorine… People died from detergents, they cleaned everything with chlorine… People ate tomatoes that were rinsed with chlorine, it was important to get rid of COVID-19.”*

“*But it is enough for a man to go out on social media and say “I am a doctor and know about the vaccine” but you don’t know…. people trust an unknown man on social media and start believing what he says. They spread things like the vaccine will kill us humans.”*

Although uncommon, some of the respondents and people they knew, who used media from countries where COVID seemed to be less widespread perceived the pandemic as mild which lowered the motivation to get vaccinated.

*“We listen to radio channels from our home country… In our home country, we do not take vaccines, we meet each other without keeping our distance and yet it has decreased… But there is a big difference in Sweden, here you are indoors, but over there (in Somalia) people are outside, sitting outside to get some air. Here you sit in small rooms.”* Somalian

### 3. Decision-making on COVID-19 vaccination

A lack of trusted information raised questions about the effectiveness, safety and possible side effects of the vaccine. The respondents felt hesitant to insert an unknown hazard into their bodies but weighed these potential threats against wanting to be protected against a lethal disease and helping society combat the pandemic. Others felt pressured to get vaccinated by their employers or the schools they attended or by being denied entrance to a country they wished to visit if they were unvaccinated.

#### 3.1 Aspects that increase vaccination hesitancy

Although several participants were aware of that a lot of the circulating information was rumours or false, it still contributed to a lot of fears of known and unknown consequences of getting vaccinated. These fears were made worse by not fully understanding how a vaccine functions, how to find reliable information or why they needed to take a booster jab every sixth month. There was also a sense that official information only focused on positive effects, while they also wanted to know negative facts to make a balanced decision.

*“There are a lot of people who don’t want to get vaccinated at all and I don’t blame them. Because they don’t have information about the vaccine! No one who can explain to them or who can give them the right information about it. If you want to convince people of the benefits of the vaccine, you need a lot of information and explanations, and you have to give them time for it.”*

*“Do you have to take a new one after six months? After six months!? How many shots should you take in two or three years, it will be six shots…They say that the vaccination cannot prevent the COVID infection, but it only helps. What is the point of taking a new one every six months?”*

Some were confident in looking for and understanding official Swedish information, but still refrained from taking the vaccine since it does not prevent COVID-19 infection. Others figured that it was unnecessary for a healthy person who does not meet a lot of people and that vaccination is foremost needed among people who smoke or who are sick.

*“I don’t like taking medicines… I’m perfectly healthy and I’m not a danger to others. Smokers who have pneumonia will certainly feel worse if they get infected. They need the vaccine.”*

*“I stay at home and don’t usually go out much, I don’t socialise with others. I usually keep my distance from others.”*

Other concerns were that it might be inappropriate to give the same dose to everyone, regardless of age or ethnicity, but there was no information on this issue. Some also were suspicious that the vaccination would not provide any protection and only were promoted to make money for pharmaceutical companies.

*“One must not forget that the human body’s immune system is different. They can respond differently to the vaccination as well as the medication. You don’t prescribe medicine to someone without first knowing the correct dose… Can an 18-year-old and a 70-year-old be the same? How many milligrams a 70-year-old takes is not the same as an 18-year-old.”*

*“The vaccine does not protect. From the very beginning, I got the feeling that it was something that was good for the pharmaceutical industry, at the expense of human life. They make a lot of money from it. The whole thing is a lie.”*

#### 3.2 Aspects that increase vaccination motivation

Factors that strengthened the motivation to take the vaccine were a good understanding of both its protective functions and potential side effects from taking the vaccine. There was also a sense of societal responsibility and that it was an important duty to get vaccinated. For others, it was mandatory to get vaccinated to attend work or school. Another motivational factor was to be able to enter other countries to visit relatives or for leisure trips.

*“We were aware of the risks, but we knew it was a preventative measure against COVID. We were ok with that.”*

*“They had said that a new vaccine had arrived and that those who did not get vaccinated were not allowed to work. Then I had to take the vaccine.”*

### 4. Suggestions to improve information dissemination effectiveness

Participants suggested many forms of improvements that would be appropriate for their methods for searching and consuming information, mainly focused on producing information in many languages and increasing dissemination through social interaction and by using trusted outlets. There were also suggestions that employers, health care centres and schools should take a larger role in spreading information.

#### 4.1 Information in many languages and formats

The participants insisted that providing information in many different languages would be a decisive part of improving information dissemination, as Swedish is not everyone’s native language and even moderate speakers might misunderstand important details. The respondents pointed out that there are people who cannot read or write any language, for whom an outreach based on spoken information would be the most efficient. In this regard, official information could be disseminated on governmental or other official websites or broadcast periodically on public service radio and TV channels. This regularity meant that everyone would know when they could find information, and people from the same social groups could take turns listening and then pass it on.

*“Several different communication channels should have been used. People understand information in different ways, by looking, listening or hearing. They would have given a complete information through different communication channels.”*

*“Someone should provide complete and sufficient information that everyone could understand and thus avoid hearsay.”*

#### 4.2 Communication through social interaction

The health guides and doulas brought forth the importance of trust and that personal communication is a first step for a community to accept new information. They emphasised the pre-established foundation of trust, underscoring that personal communication with individuals who possess an understanding of various cultural backgrounds represents the initial stride in facilitating a community’s acceptance of novel information. A trustful connection also makes it easier to freely voice concerns and to get help to navigate the health care system. The doulas and health guides, who had established broad social networks before the pandemic, organised information meetings via Zoom or WhatsApp, after which the attendees would circulate the obtained information to their family, friends and neighbours. One suggestion was also that people from different origins with different native languages could spread information by household visits in immigrant-dense boroughs, illustrating the need for communication with a trusted person from a similar culture.

*“We personally went to vaccine sites, and we talked to people in our language. We can’t just come up and ask them if they’ve been vaccinated. We first have to find out if we speak the same language… Then we start talking about the vaccine and how important it is and what it means.”*

*We look up “1177” together and we see if it matches what they heard, then we read together… We taught them to use Zoom and other platforms. They couldn’t before. But now they are experts and skilled. Some cannot read because they are illiterate. I helped them share films about Sweden and COVID-19. We used to share videos on the WhatsApp group for them so they can watch and listen.”*

Another communication attempt was having specialised experts coming to the local areas and give lectures. Such events could gather up to 40-50 participants and were positively received since people perceived the information as reliable and were given time for questions. Preferably, these campaigns should take place periodically and near their residential areas since many people are tired from working long hours and do not want to travel far. It should also be conducted in other languages than Swedish or accompanied by interpreters.

*“It would be good to have planned meetings where you gathered everyone to give them information… and not just asking the people to follow the information on television or the Swedish-Somali radio. Some won’t listen at all, they don’t have the time for it, nor the energy…. They leave home tired, and they come home tired. On Saturdays and Sundays, they are busy with household chores.”*

*“We consulted doctors who came here several times, and they talked about vaccines and explained everything to many participants… They question him “But who are you”? Then he can show with his ID that he is actually a doctor… So it becomes credible.”*

*“They spread the word to others who had not taken the vaccine and they were positively influenced by this information and our lectures. At our lectures, those who had difficulty trusting the vaccine came and accepted the vaccine after we explained to them how it works.”*

*4.3 Increased role of school, work and local healthcare centres*

Schools and employers could also have an expanded role in spreading information since they usually reach everyone who is enrolled in a class or employed by a company. Additionally, the participants wished that healthcare staff would be better at understanding cultural differences and respond to insecurities communicated in different ways by people with an immigrant background. This could improve the interaction with a patient,

## Discussion

This exploratory interview study with foreign-born participants found several themes that affected a delay or refusal to take the COVID-19 vaccine. Limited health literacy in official institutions appeared to have led to failures in understanding that immigrants are hindered from accessing COVID-19 information due to language barriers and not recognising Sweden’s health agencies, and that adapted strategies directed to foreign-born audiences could have made a difference. The unsuccessful transmission contributed to a predominant use of other sources, which seemingly facilitated the spread of misinformation and created a fearful attitude towards the COVID-19 vaccine. This complicated the decision process for getting vaccinated, where worries about hazardous side effects were balanced against wanting to be protected and helpful in society. Suggestions for future outreach was the production of information in many languages and formats, increased dissemination through social interaction and trusted platforms, and an extended role of employers, schools and the local health care system.

### Limited health literacy

Not producing information in several languages promptly was perhaps the largest shortcoming in official institutions. A study on the Swedish Public Health Agency’s social media outreach (Facebook, Twitter and YouTube), up until February 2021, showed that there were tailored information directed to the elderly and risk groups based on existing diseases (19). Only three messages were directed to immigrants, and out of those one was in another language than Swedish, despite evidence that foreign-born persons to a large extent use social media to look for information (20). During the first pandemic wave, this may relate to a lack of time for translating fast-changing information or that a variety of information channels had not been put into use. However, when the vaccine was made available in December 2020, more effective methods should have been established. Our results align with other studies, showing that public health agencies’ efforts to persuade unwilling individuals to get vaccinated often is to assume that the remedy is more information, rather than understanding core causes and adapting their methods to meet these needs (21). As many people with an immigrant background display institutional mistrust (26, 27), the willingness to understand their specific concerns would benefit trust and increase comfort in getting vaccinated.

The lack of effective strategies may relate to the communication distance and few channels that connect health agencies and foreign-born populations. A sometimes-forgotten information provider are healthcare workers, who regularly interacts with patients and can capture special needs and bridge knowledge gaps (22, 23). International studies have shown that people with an immigrant background generally have a larger distrust of healthcare personnel which originates from facing negative treatment attributed to their ethnicity (24, 25), but this has not been much studied in Sweden. One smaller questionnaire study with 52 foreign-born patients illustrated that language and cultural differences contributed to a feeling of not being heard by practitioners in the Swedish primary care (26). The respondents in the current study felt that healthcare staff did not understand that there are groups in society that lack the necessary information and need reassurance, thus an opportunity to improve trust towards the vaccine was missed.

### Consequences of not using official Swedish information

Most of the participants used social media and social contacts as their primary sources of information during the COVID-19 pandemic, which is similar to migrant groups in many other countries (27, 28). The use of a multitude of sources, especially if not being able to access official Swedish information, led to an exposure to information that was both conflicting and focused on dangerous aspects of the vaccine. A study of Hispanic- and Afro-Americans illustrated similar themes, such as distrust “the government cannot be trusted to tell the truth about COVID-19,” or hearsay “People who take a COVID-19 vaccine will be like human guinea pigs” (25). Social media has been highlighted as a concern during the pandemic (29) since this fast information outlet can play a decisive role in the perception of a vaccine. An explorative study mapping out search results on the incorrect association between the measles, mumps and rubella (MMR) vaccine and autism when using the search engine Google, showed that only 51% of the suggested web pages provided the correct information (30). However, few studies evaluate to what extent inaccurate information can overturn a vaccination decision, instead it seems to be those who are already hesitant who search for sources other than official public health agencies and vaccination providers (31, 32).

### Decision-making on COVID-19 vaccination

The pandemic itself was a new and incomprehensible situation. The lack of access to scientifically proven knowledge and an inability to find answers to their inquiries, led to further doubts about the vaccine, which seemed hard to overcome. This has been noted before in controversies surrounding a vaccine; once distrust is established, reinstalling trust can be difficult (18). Vaccination acceptance is a sociological macroprocess influenced by the close social network and societal pressures to perform a duty, as well as a historical and sociocultural context, and relates to previous experiences of vaccinations and trust in the system that delivers the vaccine (21). All these positions were observed as the respondents were affected by negative perceptions in their close social networks and lack of trust in the system, but also wanted to fulfil a societal responsibility by getting vaccinated.

### Suggestions to improve information dissemination effectiveness

The lack of access to information among people with an immigrant background in the event of national emergencies is a well-known phenomenon (33). Societies would benefit from establishing platforms that could be quickly put into use and designed to reach those who look for and consume information in other ways than the majority of the society. Most suggestions put forward among respondents in this study revolved around the importance of information in several languages, at least in the largest language groups in Sweden, and spreading information through social interaction with trusted persons and platforms. It was also suggested that local events with invited specialists should be organised, assisted by someone who could interpret and with ample time set aside for questions. Workplaces and schools could also have an extended role in providing information; in this context, there could be colleagues and other students who can be consulted about unclear aspects. Although not a direct result of this study, it was communicated to us that in many cultures important information is mostly transferred via verbal communication in the local network, especially from elderly or religious leaders, and that Swedish institutions may overestimate the power of written information such as websites or leaflets.

An EU-funded consortium COronavirus Vulnerabilities and INFOrmation dynamics Research and Modelling (COVINFORM) has frequently highlighted immigrants’ vulnerability in regard to a lack of access to, or poorly adapted, information (25). One of the consortium’s member groups, the Birmingham Local Outbreak Engagement Board, have reported trials of delivering inclusive and accessible COVID-19 communication to the diverse city of Birmingham (UK), home to 187 different nationalities (34). The National Health Services (NHS) of Birmingham and Birmingham Public Health Division used an intensive information campaign, establishing multiple information channels through which they disseminated large email and text message dispatches to local organisations, community centres, faith settings, schools and councillors. As many immigrants themselves or their family members regularly attend faith meetings, they also established meetings and virtual briefing sessions with faith institutions, ministers and pastors. Additionally, they also held bimonthly webinars and produced radio advertising to spread important information about COVID-19 and available apps provided by NHS-Birmingham. Although it was not possible to evaluate the direct effects on vaccination rates, the high user engagement on their platforms suggests that their efforts to provide the citizens with information was at least partially successful. This stands in contrast to a similar attempt of information dissemination in Norway (35), aimed at the Somali community, where the outreach was less successful due to not using established and trusted communication channels.

### Limitations and strengths

Several themes were found that are specific to having a foreign background, foremost related to language barriers and less knowledge of Swedish health care and public health institutions. However, we have not interviewed native Swedes who displayed vaccination hesitancy and, cannot elucidate if similar aspects, such as being exposed to misinformation, increase hesitancy in foreign-born and native-born citizens in Sweden alike. Because of the qualitative design, external validity cannot be tested, although this is rarely the aim of qualitative studies, and other qualitative studies have illustrated similar findings (25, 26, 33). Despite limitations in generalisability, the qualitative design is an advantage when exploring new perspectives since it facilitates the possibility of capturing nuances and delving into new themes with follow-up questions.

This study includes participants from sociodemographic groups who tend to not take part in scientific studies. This was possible because of the social networks of the two health guides, who also could conduct two of the focus groups in the participants’ native language. Previous studies have pointed out that studies among foreign-born tend to be conducted in Swedish, which requires good language skills and excludes the most vulnerable persons in terms of living conditions and health (34).. The focus groups independently described similar themes and the stability of these responses indicates that our findings have transferability to other similar populations. We also recruited doulas and other health guides who due to the size of their catchment areas and the timing of the interviews (spring 2022), could provide an overview of various themes, and had experience of information dissemination efforts and the effectiveness of these efforts.

## Conclusions

Vaccine hesitancy, a barrier to full population inoculation against highly infectious diseases, is more common in some sociodemographic groups. In this study, insufficient health literacy on both the senders’ and receivers’ end, meant that important information about the COVID-19 vaccine did not reach everybody equally. This led to an increased exposure to misinformation and diminishing the chances of making well-informed decisions on vaccination. Suggestions for improved information outreach focused on the use of already established and trusted communication channels and the dissemination of information in several languages.

## Ethical approval

This study has been approved by the Swedish Ethical Review Authority (EPM dnr 2021-00177, EPM dnr 2022-02942-02).

## Data Availability

Data are not publicly available. Anonymized data are available upon reasonable request from the first author and pending fulfilment of legal requirements.

## Acknowledgements

We would like to thank Saido Mohamed for her help in recruiting and interviewing participants and contributions with her cultural knowledge.

